# Serum Lactate–Based Stratification for Seizure Diagnosis in Resource-Limited Neurologic Emergency Settings

**DOI:** 10.1101/2025.05.01.25326838

**Authors:** Shin-ichiro Osawa, Satoshi Miyata, Kensuke Fujita, Tsuyoshi Kawamura, Ichiro Suzuki, Kensuke Kimura, Toshimi Okushima, Akihide Konn, Hidenori Endo

## Abstract

**Background:** Epileptic seizures (SZ) and postictal neurological deficits are common stroke mimics that challenge acute clinical decision-making, especially in emergency settings with limited diagnostic resources. MRI or EEG is often unavailable at initial presentation. We aimed to determine whether routinely available serum lactate levels could distinguish SZ from stroke based on information accessible immediately on hospital arrival.

**Methods:** We retrospectively reviewed 22,430 consecutive emergency department visits and analyzed 637 patients transferred by ambulance with suspected neurologic emergencies identified via keyword screening at the regional emergency call center. All patients underwent venous blood gas analysis and non-contrast CT on arrival. We evaluated whether serum lactate and other routine laboratory variables (pH, actual base excess, glucose, WBC count, platelet count, and PT-INR) correlated with final neurological diagnoses. Classification and Regression Tree (CART) analysis was used to identify cutoffs predictive of SZ.

**Results:** Age, pH, lactate, and actual base excess were significantly associated with final diagnoses. Lactate levels were significantly higher in SZ and subarachnoid hemorrhage groups than in other groups. For distinguishing SZ from ischemic stroke (infarction and TIA), CART analysis yielded a serum lactate cutoff of 4.05 mmol/L. Among patients with lactate ≥4.05 mmol/L and age <59.5 years, SZ probability was 100%. A four-quadrant model combining lactate and age stratified SZ probabilities from 9.1% to 100%. The area under the ROC curve for lactate in distinguishing SZ from stroke was 0.800.

**Conclusions:** Serum lactate, when assessed upon emergency department arrival, may contribute significantly to differentiating seizures from acute ischemic stroke. A simple decision model using only lactate and age may aid in diagnostic stratification in neurologic emergencies—even in resource-limited settings where advanced imaging or EEG is not readily available.

**Registration:** None.

## Introduction

Timely recanalization therapy is central to the management of acute ischemic stroke, but its efficacy is often constrained by diagnostic uncertainty and time-sensitive decision-making requirements^1,2^. While advanced imaging techniques have improved stroke diagnosis, physicians frequently must decide on treatment eligibility based on limited information, particularly in emergency settings where rapid intervention is crucial. A major diagnostic challenge arises when epileptic seizures (SZ) and postictal neurological deficits mimic stroke symptoms, delaying appropriate recanalization therapy— especially when key tools such as magnetic resonance imaging (MRI) or electroencephalography (EEG) are not readily available^3–5^.

In such resource-limited contexts, initial assessments typically rely on non-contrast computed tomography (CT), venous blood sampling, patient history, and brief neurological examinations. Enhancing diagnostic precision using only these basic data could help guide clinical decision-making without relying on specialized modalities.

Serum lactate, a biomarker of anaerobic metabolism, has been proposed as a potentially useful marker for differentiating SZ from other acute neurologic conditions. However, its real-world diagnostic value—particularly in the acute setting of suspected neurologic emergencies—remains unclear^6^.

We hypothesized that serum lactate, when combined with minimal essential clinical data, could help distinguish SZ from ischemic stroke at the time of hospital arrival. In this study, we retrospectively analyzed a consecutive cohort of patients presenting with suspected neurologic emergencies to evaluate whether serum lactate and other routinely collected variables could support early differential diagnosis. Our goal was to develop a practical, evidence-based decision model applicable in time-critical, resource-constrained emergency settings.

## Methods

### Patients and treatment protocol

A total of 22,430 consecutive patients presented to a single comprehensive emergency and critical care center (Hachinohe City Hospital, Hachinohe, Aomori, Japan) between April 1, 2015, and March 31, 2016. Of these, we retrospectively analyzed 661 patients who had been transported by ambulance under a designated “neurological emergency code.” After excluding 24 patients due to insufficient data, a total of 637 patients were included in the final analysis (Fig. 1). The “neurological emergency code” was assigned by the Aomori Prefecture public emergency call system based on specific keywords and prompted transfer acceptance by the hospital’s emergency department. The keywords included: limb motor weakness, facial asymmetry, speech difficulty (including dysarthria and aphasia), impaired consciousness without overt cardiopulmonary failure, and convulsions.

**Figure 1:**
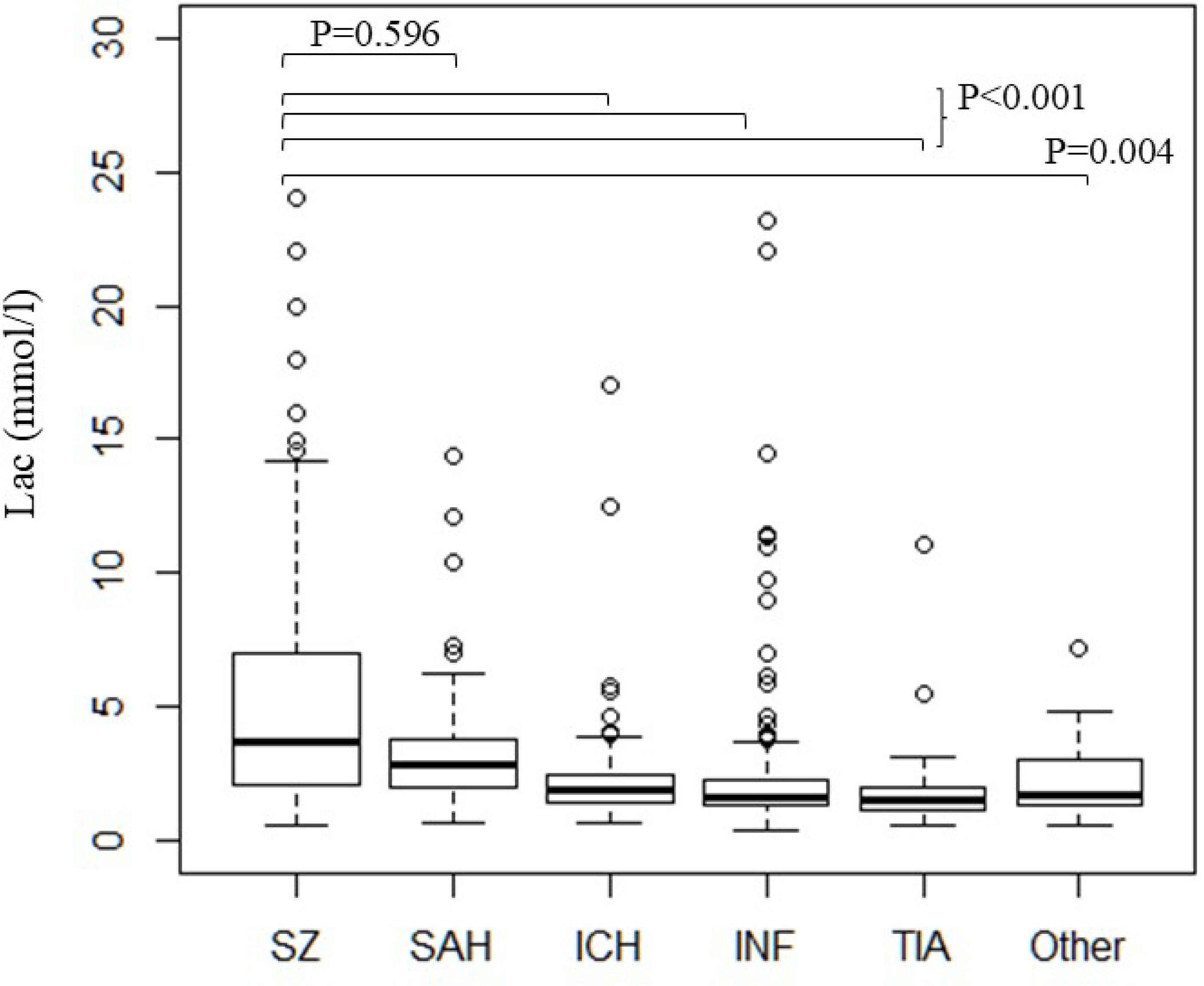
Study flow. Comparison of serum lactate level among each stroke type

All patients underwent immediate neurological evaluation, non-contrast computed tomography (CT), and venous blood sampling upon arrival to assess eligibility for intravenous thrombolytic therapy for acute ischemic stroke.

The study protocol was approved by the Hachinohe City Hospital Ethics Committee (Approval No. 1601). All patient data were anonymized before analysis.

### Data collection procedures

All patient data were collected from hospital electronic medical charts. The collected factors were as follows: sex, age, blood sample parameters of pH, lactate, actual base excess (ABE), white blood cell count (WBC), platelet, international normalized ratio of prothrombin time (PT-INR), and blood glucose. We selected these factors based on their necessity for determining the indication for intravenous thrombolytic therapy. We compared the relationship between these parameters and final diagnoses after detailed examination following admission, including neurological examination, CT (with or without contrast enhancement), MRI, EEG, and clinical findings from precise history-taking. Patients were grouped according to the final diagnoses: SZ, complete infarction (INF), transient ischemic attack (TIA), subarachnoid hemorrhage (SAH), intracerebral hemorrhage (ICH), and Other.

### Statistical analysis

Patient characteristics in each group were analyzed by analysis of variance (ANOVA). We used univariate and multivariate stepwise selection with backward elimination analysis to compare the correlation between variables available on arrival and patient groups based on final diagnoses. Independent t-tests were used to compare continuous variables. Lactate levels in each group were analyzed by a nonparametric Dunnett-type many-to-one multiple comparison procedure. A *P* value of less than 0.05 was considered to indicate a significant difference.

The sensitivity and specificity of lactate levels for distinguishing between SZ and ischemic stroke (INF and TIA) were analyzed using a receiver operating characteristics curve. Classification Analysis of Regression Tree (CART) analysis was performed to identify the cutoff value of the parameters significantly correlated with the diagnosis group in univariate analysis.^7,8^

## Results

We obtained baseline characteristics for all 637 patients (Table 1). Overall, we found a significant difference in several factors among the patients in the final diagnosis groups. Significant differences in WBC, pH, lactate levels, and ABE between groups were detected using ANOVA (*P* < 0.001 for all). In the SZ group, patients were younger with higher lactate levels and lower ABE levels. In the SAH group, patients had higher WBC counts, higher glucose and lactate levels, and lower ABE levels. In the INF group, patients were older. Among the parameters, we analyzed the lactate level between all groups by inter-group comparison. The lactate level was significantly higher in the SZ group than in the ICH, INF, TIA, and Other groups (*P* < 0.001, *P* < 0.001, *P* < 0.001, and *P* = 0.004, respectively), but did not differ significantly from that in the SAH group (*P* = 0.596) (Fig. 2, Supplementary table 1). In the supplementary analysis excluding the non-stroke group, the lactate level was significantly higher in the SAH group compared with the other groups (Supplementary table 2). Next, we selected the SZ, INF, and TIA groups as candidates for differential diagnosis using lactate, as SAH and ICH could be identified as hemorrhagic through rapid imaging modalities such as CT. Univariate analysis of variable parameters between the SZ group and the INF and TIA groups revealed a significant association of differential diagnoses with five parameters: age, WBC, pH, lactate, and ABE (*P* < 0.001, *P* = 0.039, *P* < 0.001, *P* < 0.001, *P* < 0.001, respectively). The following multivariate logistic regression analysis demonstrated that age and serum lactate level were significant predictors of SZ (odds ratio [95% confidence interval, 1.063 (1.047-1.081)], P<0.001; and [95% confidence interval, 0.744 (0.671 - 0.814)], P<0.001, respectively) (Table 2). The reliability of lactate levels for differentiating between the SZ group and the INF and TIA groups was assessed using a receiver operating characteristics (ROC) curve, with an area under the curve of 0.800 (0.725–0.895) (Fig. 3). Classification and Regression Tree (CART) analysis identified a serum lactate cutoff of 4.05 mmol/L as the primary discriminator for SZ diagnosis. Subsequent stratification used age thresholds of 59.5 years (for lactate ≥4.05 mmol/L) and 39.5 years (for lactate <4.05 mmol/L). The probabilities of SZ diagnosis in each subgroup were as follows:

- 100% in patients with lactate ≥4.05 mmol/L and age <59.5 years,
- 84.6% with lactate <4.05 mmol/L and age <39.5 years,
- 48.4% with lactate ≥4.05 mmol/L and age ≥59.5 years, and
- 9.1% with lactate <4.05 mmol/L and age ≥59.5 years (Fig. 4).

**Figure 2:**
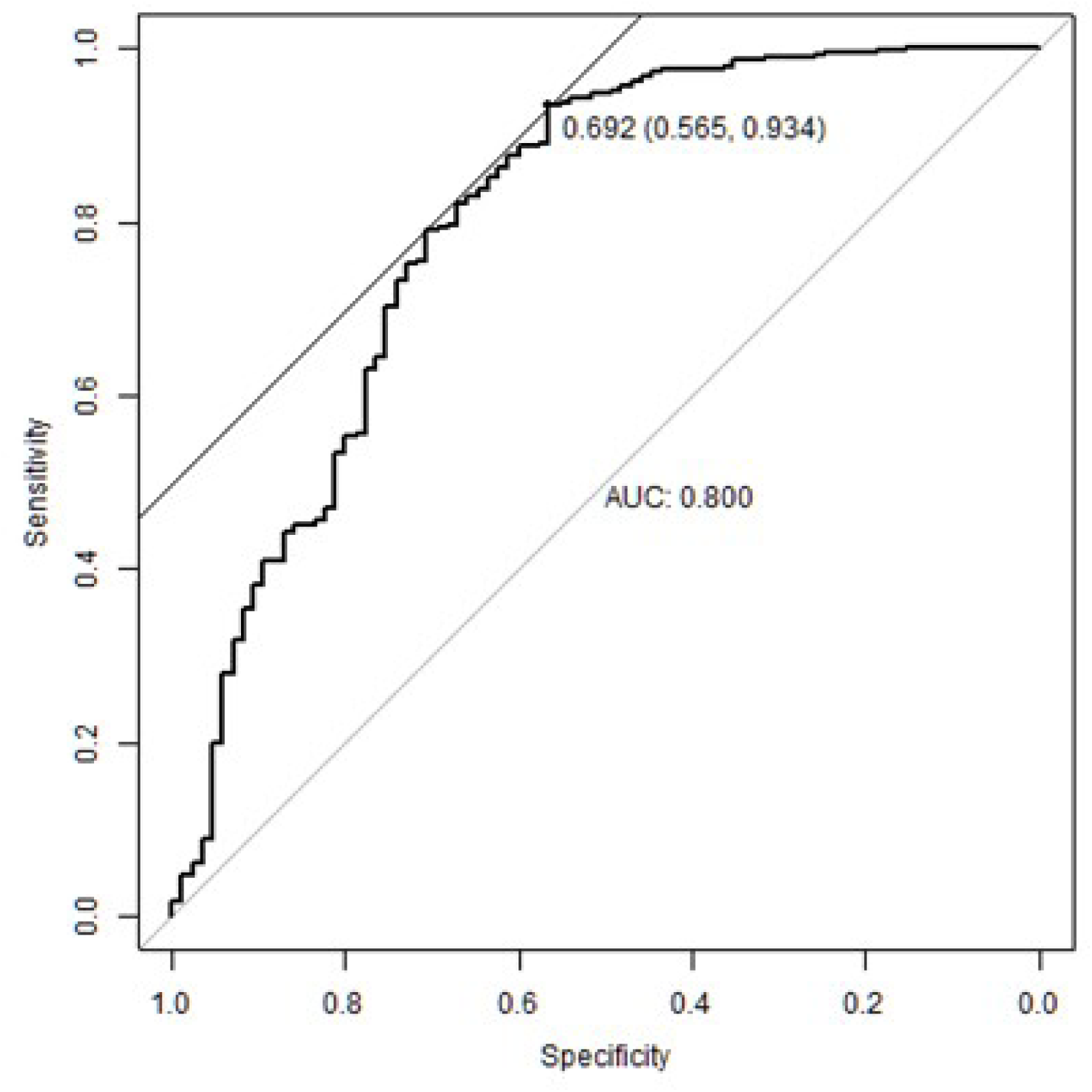
Comparison of serum lactate level among neurologic emergency types Inter-group comparisons were analyzed using ANOVA, while comparisons between individual groups were performed using the nonparametric Dunnett-type many-to-one multiple comparison procedure. The ROC curve between SZ vs. (INF+TIA)

**Figure 3:**
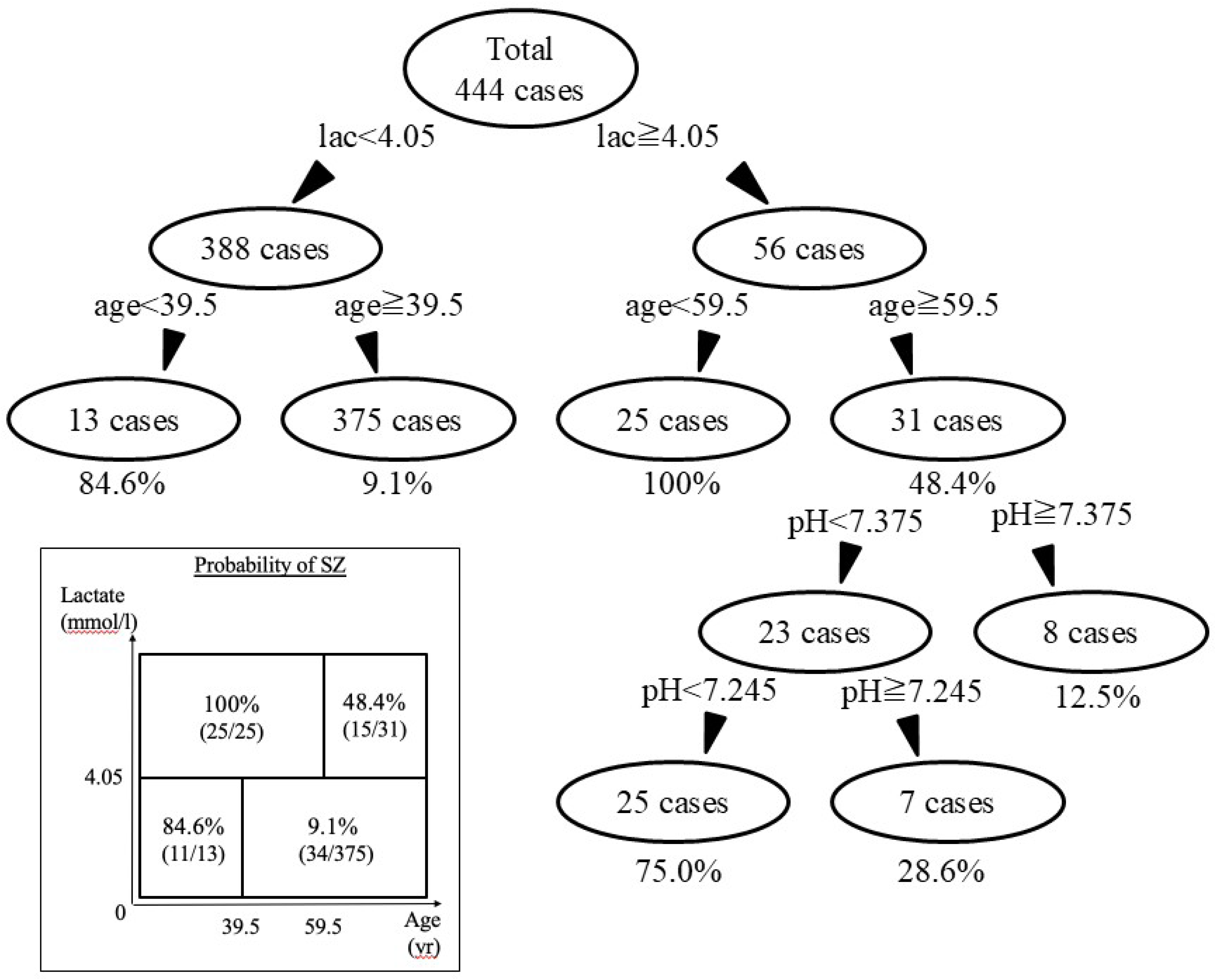
The ROC curve between SZ vs INF+TIA ROC = receiver-operating characteristic

**Figure 4:**
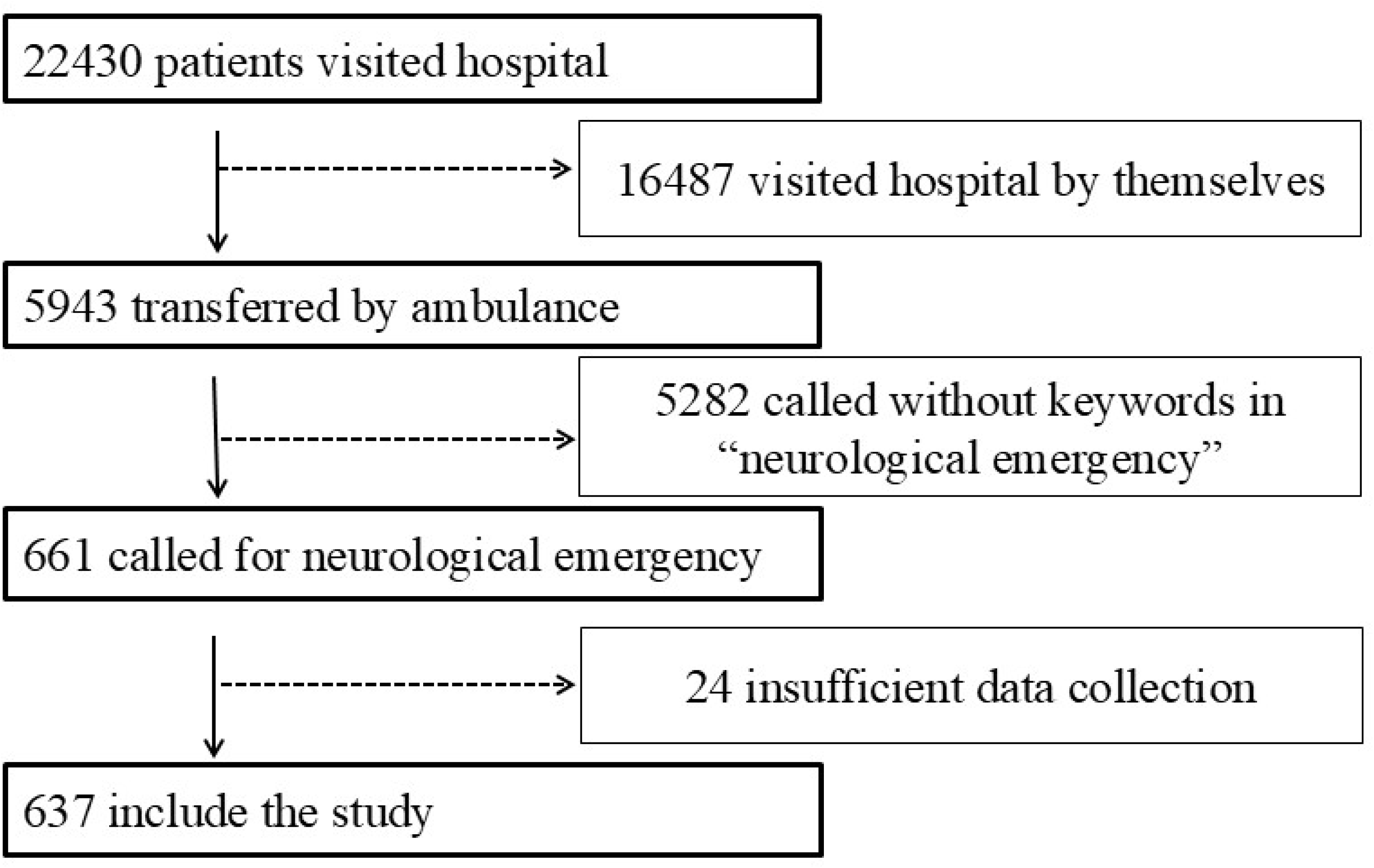
CART analysis for seizure probability. (inset) Diagnostic model of SZ probability CART = classification and regression tree; SZ = epileptic seizure. yr=years.

**Table 1:** Characteristics of the participants. All data are presented as mean ± SD, unless stated otherwise. Statistical significance was analyzed using ANOVA. WBC = white blood cell, PT-INR = international normalized ratio of prothrombin time, ABE = actual base excess, SZ = epileptic seizure, SAH = subarachnoid hemorrhage,. ICH = intracerebral hemorrhage, INF = infarction, TIA = transient ischemic attack.

|  | All<br>(n=637) | SZ<br>(n=85) | SAH<br>(n=47) | ICH<br>(n=121) | INF<br>(n=313) | TIA<br>(n=46) | Other<br>(n=25) | p-value |
| --- | --- | --- | --- | --- | --- | --- | --- | --- |
| Sex (male) | 357 (56%) | 55 (64.7%) | 18 (38.3%) | 73 (60.3%) | 177 (56.5%) | 21 (45.7%) | 13 (52%) | 0.039 |
| Age (year old) | 69.74 ±<br>16.07 | 56.87 ± 22.55 | 63.43 ± 14.37 | 69.7 ± 12.48 | 74.94 ± 12.57 | 69.59 ± 13.71 | 60.84 ± 19.55 | < 0.001 |
| WBC (x10<sup>3</sup>/μl) | 8.36 ± 3.76 | 8.59 ± 3.81 | 11.43 ± 4.13 | 9.27 ± 4.35 | 7.67 ± 3.34 | 6.73 ± 2.08 | 8.93 ± 2.97 | < 0.001 |
| Platelet (x10<sup>4</sup>/μl) | 21.29 ± 7.68 | 23.17 ± 9.65 | 22.73 ± 7.28 | 19.59 ± 5.89 | 21.08 ± 7.93 | 21.95 ± 5.78 | 21.84 ± 6.88 | 0.019 |
| PT-INR | 1.12 ± 0.66 | 1.13 ± 0.31 | 1.23 ± 1.45 | 1.2 ± 1.02 | 1.08 ± 0.34 | 1.09 ± 0.33 | 1.12 ± 0.35 | 0.477 |
| Glucose (mg/dl) | 148.69 ±<br>65.57 | 150.79 ± 87.03 | 186.23 ± 66.13 | 149.41 ± 60.5 | 145.15 ± 61.39 | 132.2 ± 38.9 | 141.41 ±<br>72.75 | 0.001 |
| pH | 7.38 ± 0.09 | 7.33 ± 0.12 | 7.34 ± 0.16 | 7.37 ± 0.09 | 7.40 ± 0.05 | 7.39 ± 0.06 | 7.39 ± 0.06 | < 0.001 |
| Lactate (mg/dl) | 2.72 ± 3.02 | 5.62 ± 5.16 | 3.53 ± 2.8 | 2.27 ± 1.92 | 2.13 ± 2.29 | 1.85 ± 1.62 | 2.25 ± 1.52 | < 0.001 |
| ABE | 0.09 ± 4.66 | -3.26 ± 7.28 | -2.14 ± 6.21 | 0.52 ± 4.75 | 1 ± 2.84 | 1.05 ± 3.31 | 0.66 ± 3.49 | < 0.001 |

WBC: white blood cell; PT-INR: international normalized ratio of prothorombin time; ABE: actual base excess; SZ: seizure; SAH: subarachnoid hemorrhage; ICH: intracerebral hemorrhage; INF: infarction; TIA: transient ischemic attack.

**Table 2:**
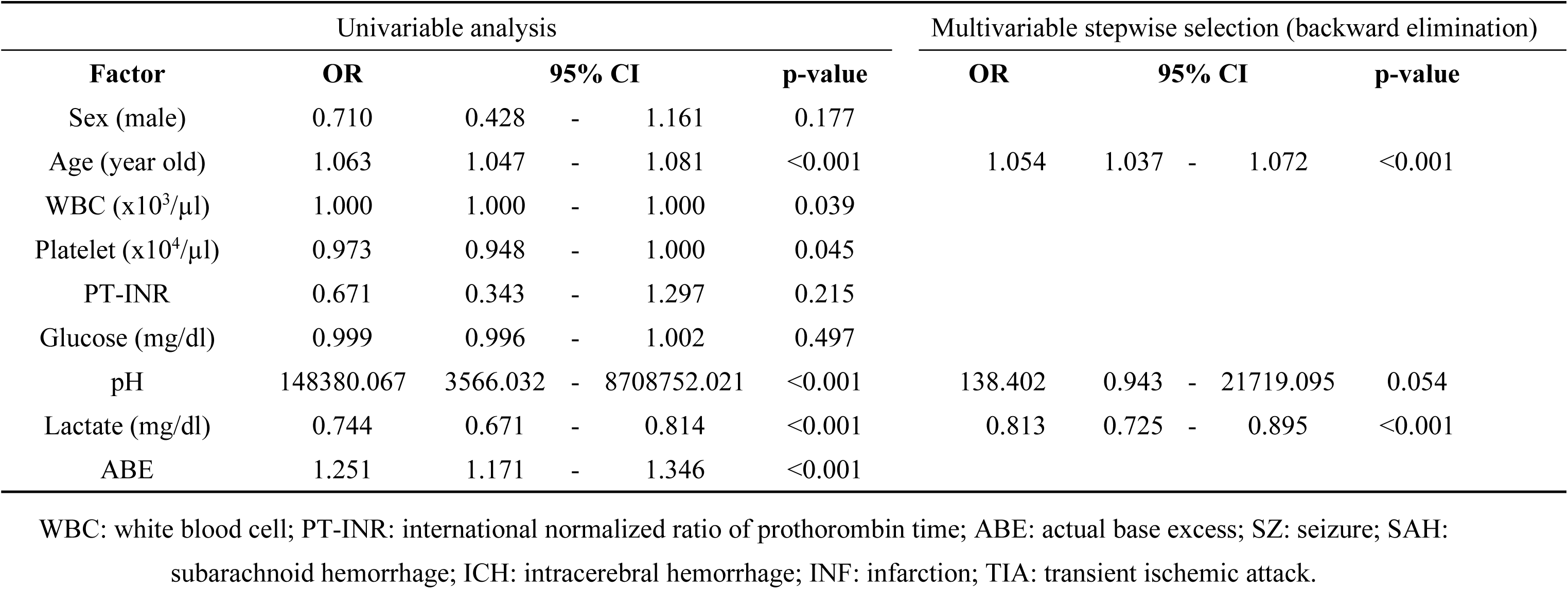
Correlations between SZ and INF + TIA groups OR = odds ratio, CI = confidence interval.

Based on these findings, we propose a tetrameric decision model combining serum lactate and age to aid in differentiating SZ from ischemic stroke (INF and TIA) in neurologic emergencies (Fig. 4, inset).

## Discussion

Here, we report data on biomarkers that may facilitate the differential diagnosis of patients with suspected neurological emergencies. Our analysis yielded two key findings. First, we established the prevalence of elevated blood lactate levels across various conditions, such as SZ, SAH, ICH, INF, and other acute neurologic conditions. Second, lactate, when combined with age, served as a strong predictor for distinguishing SZ from ischemic stroke, particularly in emergency settings where CT is available but advanced modalities are not.

### Stroke mimics

Due to the brief time window for initiating thrombolysis after symptom onset in patients with suspected acute ischemic stroke, rapid neurological examination and history-taking are essential. This urgency makes it challenging to distinguish acute stroke from stroke mimics, such as SZ, metabolic disorders, complex migraines, and psychological problems^1,2,6,9,10^. Stroke mimics account for up to 20% of patients evaluated under stroke protocols^9,11^. SZ is the third most frequent diagnosis (4.0%) among stroke mimics^12^ and is frequently difficult to identify due to its variable presentations and sometimes misleading perfusion imaging findings, such as peri-ictal hyperperfusion^13^. SZ can also be the presenting symptom of a true stroke, with 1.2% of ischemic strokes occurring within 72 hours of onset^14^. Yet the perfusion patterns of SZ and stroke frequently overlap, and postictal paresis or negative motor phenomena may mimic stroke-related neurological deficits^15,16^. Thus, improving differentiation tools is crucial for appropriate patient selection and timing of recanalization therapies.

In a retrospective analysis of 95 patients presenting with convulsions, 45 were finally diagnosed with SZ, and 50 were diagnosed with acute ischemic stroke^12^. Lucas *et al.* reported that 40% of patients with acute ischemic stroke and 73% of patients with SZ exhibited abnormal CT perfusion findings. In the group experiencing SZ, hyperperfusion was observed more frequently (36% compared to 2% for acute ischemic stroke), and the diagnosis of SZ was made with high specificity (98%) but low sensitivity (35%)^13^. In 24% of patients in the seizure group and 2% of patients in the acute ischemic stroke group, hypoperfusion was not limited to a vascular area, occurring in 38% of cases in each group. Distinguishing the true etiology remains challenging across hypo-, hyper-, and normoperfusion patterns due to overlapping vascular-related findings^17^.

### Serum lactate as a biomarker of SZ

Lactate, a biomarker of anaerobic metabolism, is typically measured to assess the severity of the illness or overall health. Serum lactate is used in case-control studies to assess the severity of acute ischemic stroke^6^. Patients with generalized tonic-clonic seizures have higher serum lactate levels^11,18^. In case-control trials, it is useful as a diagnostic marker for patients with suspected SZ^10^. Neither the predictive value of lactate nor the true distribution of serum lactate levels in suspected neurological emergencies, such as stroke, seizures, and others, however, have been thoroughly examined. In this study, we used less-biased admission data from consecutive patients. On admission, patients with SZ and SAH had significantly higher serum lactate levels than patients with any other kind of neurological emergency. Additionally, we reported the actual prevalence of serum lactate levels in all suspected neurological emergency patients, providing reference data for future studies.

As it is considerably simpler to rule out intracranial hemorrhagic strokes like SAH or ICH with CT, our findings highlight the predictive value of serum lactate levels in distinguishing SZ from ischemic stroke, as demonstrated by ROC curve and CART analysis.

We demonstrated that, particularly through CART analyses, that a serum lactate level of 4.05 mg/ml serves as the primary discriminating factor for SZ diagnosis. Age serves as a second differentiating point, with age cutoff values of 39.5 years in the lower lactate group and 59.5 years in the higher lactate group. With 100% (25/25) of patients in the category of “lactate ≥4.05 mg/ml and age <59.5years” and 84.6% (11/13) of patients in the category of “lactate <4.05 mg/ml and age <39.5 years”, respectively, each two-staged cutoff model predicts a higher chance of SZ diagnosis. Based on these findings, an age-matched serum lactate score may provide further information about the value of serum lactate level as a diagnostic marker in neurological emergencies. This approach can be used for patients experiencing seizures without a convulsion.

We considered several factors contributing to these results. One key factor is the epidemiological perspective. Patients with SZ were significantly younger than those with INF and TIA, consistent with the known characteristics of epilepsy and seizures, which occur in people of all ages^19,20^, whereas ischemic stroke is a vascular disease that occurs more frequently in older individuals^21^.

The second is the overall effect of the condition on the production and removal of lactate. The biggest source of lactate production, skeletal muscle volume^22^, varies with sex^23^.

Given the costs and potentially fatal risks associated with an incorrect diagnosis, including invasive procedures and unnecessary medications, lactate may serve as a readily available and life-saving laboratory test for this specific patient population.

As stroke treatment tactics have advanced, doctors are able to make rapid decisions about the best course of action, particularly when treating suspected acute ischemic stroke^24^. The efficacy of these approaches and tools is supported by the data, but not all stroke centers can use them due to the unavailability of specialist MRI or contrast CT equipment for tissue-based recanalization therapeutic indications ^25,26^.

As of right now, no postictal laboratory test can conclusively confirm or rule out an epileptic seizure. Prolactin^27^ and ammonia^28^ have demonstrated utility in distinguishing psychogenic non-epileptic seizures from SZ. While routine use in general clinical practice remains challenging, future studies may prove these markers beneficial.

### Limitations

The study has several limitations. First, it was a retrospective, single-center analysis conducted in Japan, and the study population consisted almost entirely of Asian patients. Although the cohort was consecutive with minimal exclusions, selection bias cannot be entirely excluded. Second, the groups might have had overlapping conditions. For example, hemorrhagic stroke occasionally presents with convulsions but was not included in the SZ group in this study. Third, data on the time-course from seizure to arrival was lacking. The serum lactate dynamics are considered fast and complicated^29^, this lack of temporal resolution may have affected the interpretation of serum lactate levels.

Further prospective, multi-center studies are needed to validate our findings and assess generalizability across broader populations and clinical settings.

### Conclusions

A stepwise classification model based on serum lactate level and patient age at arrival demonstrated strong diagnostic accuracy in distinguishing seizures from ischemic stroke. When combined with minimal imaging, this approach may support rapid and informed decision-making in neurologic emergencies, particularly in settings without access to advanced diagnostics.

### Contributors

SO developed the hypothesis and conceived the study. SO, HE, KN, KK, TO, TK and AK compiled and analyzed the clinical data. TM analyzed the data statistically and confirmed the accuracy. All authors critically revised the first draft and final manuscript.

## Data Availability

Data not provided in the article may be shared (anonymized) at the request of any qualified investigator for the purposes of replicating the procedures and results. To ensure participant confidentiality, the conditions of our ethics approval do not permit public archiving of study data. Readers seeking access to the data should contact the corresponding author (S.O.).

## Acknowledgement

We would like to thank all participants for HACHInohe Neuro-Endovascular Treatment (HACHI-NET) project. We thank Ms. Michiyo Sakuta for her contribution to data collection.

---

Supplementary table 1: Comparison of serum lactate level between SZ and stroke types SZ = epileptic seizure, SAH = subarachnoid hemorrhage, ICH = intracerebral hemorrhage, INF = infarction, TIA = transient ischemic attack.

Supplementary table 2: Correlation between SAH and stroke types, excluding SZ SAH = subarachnoid hemorrhage, ICH = intracerebral hemorrhage, INF = infarction, TIA = transient ischemic attack.

